# The Role of Sociodemographic and Psychosocial Variables in Early Childhood Development: A Secondary Data Analysis of the 2014 Multiple Indicator Cluster Survey in the Dominican Republic

**DOI:** 10.1101/2021.02.22.21251784

**Authors:** Laura V. Sánchez-Vincitore, Arachu Castro

## Abstract

**Introduction:** The association between sociodemographic factors, such as poverty, lack of maternal schooling, and being male at birth, and childhood developmental delay and poor educational outcomes has been established in the Dominican Republic. However, moderating factors present or introduced in families to buffer the effects of such factors on childhood development are still unknown.

**Methods:** We conducted a secondary analysis of the 2014 Multiple Indicator Cluster Survey for the Dominican Republic, a national household survey focused on maternal and child health and development. The first aim of our study was to determine if a sociodemographic model predicted early childhood development. The second aim was to determine if a psychosocial model (including family childrearing practices, discipline, and early childhood stimulation) predicted early childhood development above and beyond the sociodemographic model.

**Results:** We found that both models predicted childhood development significantly, but that the psychosocial model explained 5% more variance than the sociodemographic model. The most relevant sociodemographic predictors were socioeconomic position and mother’s education, which uniquely explained 21% and 17% of the early childhood development variance, respectively. The most salient psychosocial predictors of early childhood development were: 1) attendance to an early childhood education program, which uniquely explained 15.0% of the variance; 2) negative discipline, which uniquely explained 12.4% (negative impact); 3) the number of children’s books at home, which uniquely explained 12.0%; and 4) stimulating activities at home, which uniquely explained 5%.

**Conclusion:** These results have multiple implications for social programs that aim to improve children’s developmental potential in contexts of poverty. Although the results show a protective effect of psychosocial factors, sustainable and large-scale intervention should not be limited to just buffering effects, but to actually solve the underlying problem which is that poverty prevents children from reaching their developmental potential.

## Introduction

Early childhood development (ECD) is a global health concern, particularly in contexts of poverty (1-4). Approximately 249 million children in lower- and middle-income countries might not develop fully due to poverty, inequality, violence, stress, deficient care, lack of education and opportunities, among others (5). There is a 10% prevalence of cognitive delay in preschoolers in lower and middle-income countries, which calls for evidence-based prevention strategies to avoid the loss of developmental potential (6).

The Dominican Republic is no exception, facing multiple challenges regarding childhood development and other educational outcomes, partly due to adverse early childhood experiences. According to the most recent multidimensional childhood poverty index, 49.3% of Dominican children lived in poverty and 7.7% lived in extreme poverty in 2012 (7); neonatal and under-five mortality rates are high, with 19 and 29 deaths per 1,000 live births, respectively, in 2018 (8); and 21.5% of newborns were born to an adolescent mother in 2019, the highest percentage in Latin America and the Caribbean (9). Additionally, sociocultural childrearing practices sometimes promote psychological aggression and physical punishment to discipline children, evidenced by reports stating that 62.9% of Dominican children aged 1 to 14 years have experienced such a method of discipline regardless of socioeconomic position (10). Results of numerous studies have demonstrated that negative experiences in childhood may alter children’s development and brain chemistry (3, 11-19), and these effects are detectable starting in early childhood (20). Impairments to brain development carry implications for cognitive, social, and behavioral deficits that may affect children as they enter adolescence and adulthood (3).

Children’s stress responses to these adverse exposures have been characterized as positive, tolerable, or toxic, depending on their intensity and duration and their potential to impact physiologic regulation (21). Mild to moderate adverse exposures can trigger brief and positive stress responses that can promote growth, particularly in the presence of a supportive caregiver (3). In the absence of protection from supportive caregivers, the exposure to more adverse circumstances can lead to toxic stress—that is, excessive or prolonged activation of biological stress responses that can alter brain structure and function, and lead to impaired linguistic, cognitive, and social-emotional skills, as well as a life-long greater risk for chronic disease (3, 22).

Children who live in poverty are more likely to be exposed to forms of deprivation such as neglect, economic hardship, and undernutrition, to threats such as abuse, caregiver substance use, violence from or among caregivers, community violence, as well as to other forms of early adverse experience, than children from more affluent, supportive households (1-4). The correlation between socioeconomic position and childhood development exists throughout the world, including high-, low-, middle- (23, 24), and very low-income countries (23). Experiencing poverty during childhood can also impact the development of selective attention—that is, the ability to focus on relevant stimuli while ignoring irrelevant ones (3, 25-29). Immature selective attention triggers a cascade of negative effects in developing other complex educational skills, such as language, literacy, and math (30). Selective attention is a malleable skill sensitive to training in early human development, both directly (31) and indirectly through family-based interventions (32). Grantham-McGregor and colleagues described two pathways in which poverty affects childhood development and school achievement (33). The first pathway, related to the lack of food, hygiene, and overall health in poor settings, can lead to stunting, which can cause developmental delay and poor school achievement. The second pathway, which represents the effects of poverty on caregivers, such as stress, depression, low responsivity, and low education, can lead to inadequate care—which can cause stunting—and limited stimulation at home—which affects both childhood development and school achievement (33).

Multiple factors can buffer the pervasive effects of poverty on ECD, as development is sensitive to the context in which children live (34). A nurturing environment during childhood can improve brain function, protect against disease, and lead to good health during adulthood (20, 35, 36). A study conducted in 28 developing countries found a positive correlation between positive caregiving and the human development index (HDI) (37). Frongillo and colleagues found that family care behavior moderated the effects of socioeconomic position on children’s literacy and numeracy skills (38). On the other hand, negative caregiving practices in the form of physical punishment have a detrimental effect in childhood development (39) by increasing the risk of childhood anxiety (40) and behavioral problems (41). Regarding the availability of stimulating objects at home, a study conducted in 42 countries found that the number of books at home (family scholarly culture) correlates positively with academic achievement, even when controlling for parents’ education, occupation, and family socioeconomic position (42). The study showed stronger effects at the lower end of the family scholarly culture variable; that is, the effect of adding extra books on academic achievement was larger when there were fewer books at home. In addition, literacy and numeracy are associated with the home literacy environment and the availability of stimulating activities at home (38).

Providing a nurturing environment can be challenging in settings of poverty and high social inequality (36). Governments, international cooperation agencies, non-governmental organizations, and others offer intervention programs for children at risk to overcome this difficulty. The most effective interventions are those that provide service directly to children and support parents with their childrearing practices by not offering both information and building parenting skills (43, 44). Home visits programs have shown to be effective in improving the quality of the home environment, in increasing the number of playing activities with children at home (45), and in attaining moderate to large gains in social-emotional and language development (46).

Although the association between poverty and developmental delay in early childhood has been established in the Dominican Republic (7), the role of moderating factors that may be present in families of any socioeconomic position is unknown. Therefore, this study aimed to understand the extent to which the availability of positive environment and childhood stimulation buffers the effects of poverty on childhood development among Dominican children. To achieve this goal, we conducted a secondary analysis of the 2014 Multiple Indicator Cluster Survey Round 5 (MICS5) for the Dominican Republic (10), a national household survey focused on maternal and child health and development. Our study’s first aim was to determine if a sociodemographic model that accounts for demographic factors that are well known to affect childhood development—such as wealth, mother’s education, sex, and age—can predict early childhood development. The second aim was to determine if a psychosocial model that includes social factors that result from the social interaction between children, their caregivers, and the environment can predict ECD above and beyond the sociodemographic model.

## Methods

We conducted a secondary analysis of the Dominican MICS5 2014 databases. MICS is a household survey administered by local governments from low- and middle-income countries assisted by UNICEF. We selected 7,356 cases from the Dominican MICS5 databases. The cases consisted of children between 36 to 59 months of age (50% were girls). To obtain sociodemographic and psychosocial information from the participants, the following inclusion criteria had to be met: 1) caregivers had completed the MICS Under Five Children’s questionnaire modules (child’s information panel module and early childhood development module); 2) caregivers had completed the MICS Household questionnaire modules (selection of one child for child discipline module and household characteristics module); and 3) the child for which there were data on ECD on the Under Five Children’s questionnaire matched the selected child for the discipline module of the Household questionnaire.

The instrument for this study was the Multiple Indicator Cluster Survey. The variables used are included in **Table 1**. This study used de-identified secondary data that are publicly available and that cannot be re-identified. Therefore, it poses no violation of confidentiality. The authors obtained permission from UNICEF to download and use the database for research purposes. First, we conducted Pearson’s correlation analyses to evaluate the relationship between family socioeconomic position and ECD and to explore the relationship between family socioeconomic position and the other variables assessed in this study. Second, we conducted a two-step hierarchical multiple regression analysis to predict early childhood development. The first step consisted of sociodemographic factors (child’s age in months, child’s sex at birth, family socioeconomic position, and mother’s education level). The second step consisted of both sociodemographic factors and psychosocial factors (stimulating activities at home, stimulating objects at home, number of books at home, attendance to childhood program, positive discipline, and negative discipline).

**Table 1:** MICS variables used in the analysis.

| **Household questionnaire** |  |
| --- | --- |
| *Child's age* | This variable is expressed in months. |
| *Child's sex* | The scoring of this variable was 0 for girls, 1 for boys. |
| *Family socioeconomic position* | This variable categorizes families into five wealth quintiles from 1 (poorest) to 5 (richest). |
| *Mother's education level* | This variable assigns an ordinal value to maternal education (1 = none; 2 = primary; 3 = basic; 4 = upper secondary; 5 = vocational; 6 = college, university). |
| **Childhood development module (for children under five years of age)** |  |
| *Early childhood development index* | This is a series of ten yes/no statements regarding the child's observed behaviors administered to the mother or primary caregiver. Observed behaviors obtained a score of 1 and not observed behaviors obtained a score of 0. The behaviors included literacy and numeracy statements (the child identifies at least ten letters of the alphabet; the child reads at least four simple, popular words; the child knows the name and recognizes the symbol of all numbers from 1-10), physical development (the child picks up small objects with two fingers; the child is sometimes too sick to play); approaches to learning (the child follows simple directions, the child does something independently); and social and emotional development (the child gets along well with other children; the child kicks, bites or hits other children or adults; the child gets distracted easily). The answers for negative statements |
|  | (the child is sometimes too sick to play; the child kicks, bites or hits other children or adults; and the child gets distracted easily) were reversed to keep the scoring valence consistent. The childhood development index was computed by averaging all childhood development scores for each answer. The index ranged from 0 to 1. |
| *Availability of stimulating activities at home* | This instrument is a series of yes/no statements regarding developmentally appropriate activities that parents or other people do with the child. This includes reading, storytelling, singing, taking walks, playing, counting, and naming. The informer reported if the mother, father, and/or another person does each activity with the child scoring 0 for not doing the activity and 1 for doing the activity. The stimulating activities at home variable was computed by averaging the stimulating activities scores for each answer, assuming that the more activities a child does, the higher the score; and the more people doing the activities at home, the higher the score. The scores for the variable stimulating activities at home ranged from 0 to 1. |
| *Availability of stimulating objects at home* | This is a yes/no questionnaire regarding the child having access to homemade toys, store-bought toys, or stimulating objects at home. The informer reported whether the child had access to any of the three types of stimulating objects, scoring 0 for not having access, and 1 for having access. The availability of stimulating objects at home variable was computed by averaging the stimulating object scores for each answer, assuming that the more stimulating object, the higher the score. The scores from the computed variable stimulating objects at home ranged from 0 to 1. |
| *Number of books at home* | This variable represents the amounts of children's books at home. This is a numeric variable. |
| *Attendance to early childhood education center* | The variable was scored 0 for not attending and 1 for attending. This includes private or government facilities, kindergarten, or daycare. |
| **Child discipline module:** The module consists of eleven statements regarding methods of child discipline and is an adapted version from the Parent-Child Conflict Tactics Scale (47). The adult participant was asked to indicate whether they have used any of the discipline methods during the past month. For this study, we categorized the methods into two categories: positive discipline and negative discipline. |  |
| *Positive discipline* | Averaging answers to the following statements computed this variable: took away privileges, explained why the behavior was wrong, and gave the child something else to do. The positive discipline variable scores ranged from 0 to 1. |
| *Negative discipline* | Averaging answers to the following statements computed this variable: shook the child; shouted, yelled or screamed at the child; spanked, hit or slapped child on the bottom with bare hands; hit the child on the bottom or elsewhere with a belt, brush, stick, etc.; called child dumb, lazy or another name; hit or slapped the child on the face, head or ears; hit or slapped the child on the hand, arm or leg; and beat child up as hard as one could. The negative discipline variable ranged from 0 to 1. |

## Results

We ran internal consistency analyses on the ECD index, positive discipline, and negative discipline. We found that the ECD index had poor internal consistency (α = .5) and that both positive and negative discipline had acceptable levels of internal consistency (α = .6).

We conducted a Pearson’s correlation analysis to evaluate the relationship between ECD and family socioeconomic position. We found a significant positive correlation between the two variables, indicating that the higher the family socioeconomic position, the higher the childhood development score (*r*(7354) = .29, *p* <001). However, the magnitude of the correlation was small (*r*^*2*^ = .08). **Table 2** contains descriptive statistics.

**Table 2:**
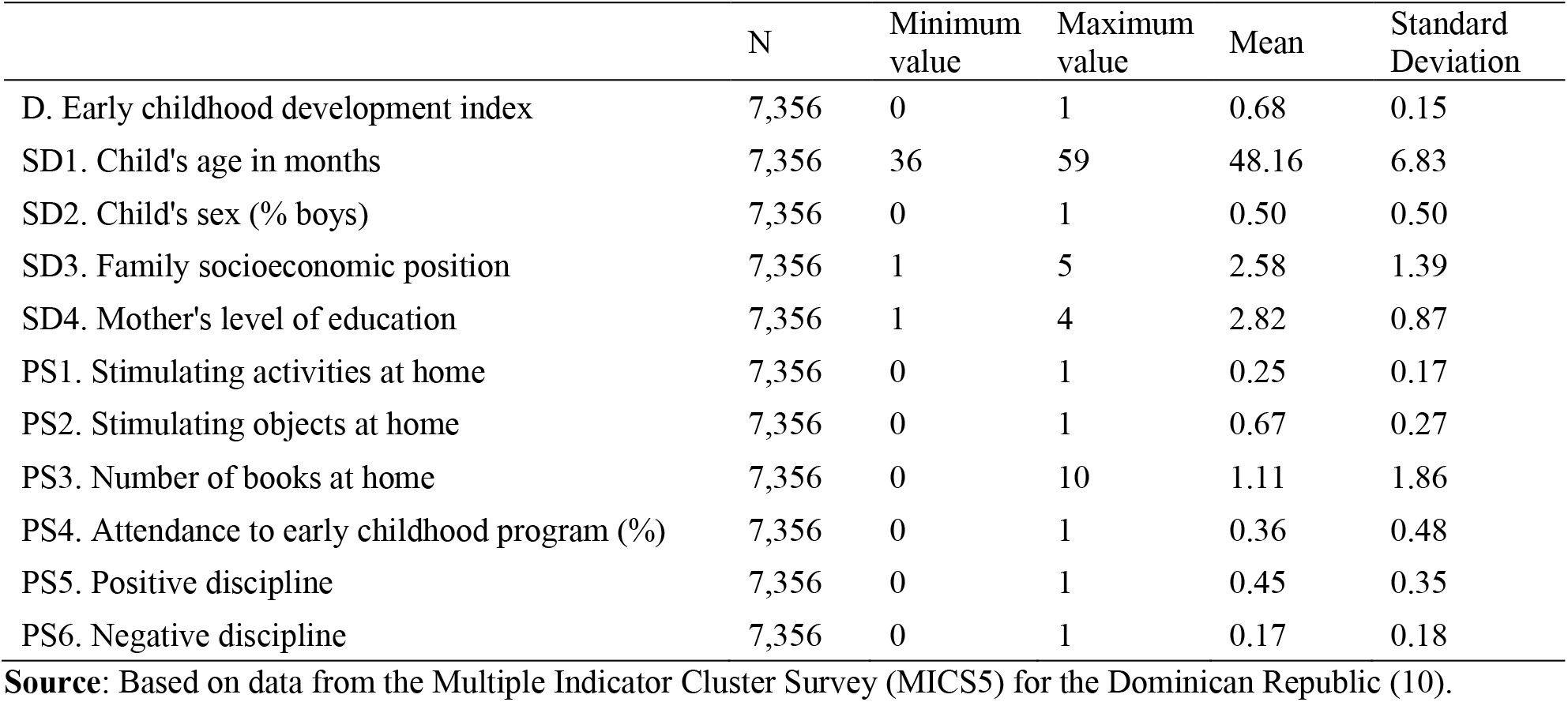
Descriptive statistics for tested variables.

|  | N | Minimum value | Maximum value | Mean | Standard Deviation |
| --- | --- | --- | --- | --- | --- |
| D. Early childhood development index | 7,356 | 0 | 1 | 0.68 | 0.15 |
| SD1. Child's age in months | 7,356 | 36 | 59 | 48.16 | 6.83 |
| SD2. Child's sex (% boys) | 7,356 | 0 | 1 | 0.50 | 0.50 |
| SD3. Family socioeconomic position | 7,356 | 1 | 5 | 2.58 | 1.39 |
| SD4. Mother's level of education | 7,356 | 1 | 4 | 2.82 | 0.87 |
| PS1. Stimulating activities at home | 7,356 | 0 | 1 | 0.25 | 0.17 |
| PS2. Stimulating objects at home | 7,356 | 0 | 1 | 0.67 | 0.27 |
| PS3. Number of books at home | 7,356 | 0 | 10 | 1.11 | 1.86 |
| PS4. Attendance to early childhood program (%) | 7,356 | 0 | 1 | 0.36 | 0.48 |
| PS5. Positive discipline | 7,356 | 0 | 1 | 0.45 | 0.35 |
| PS6. Negative discipline | 7,356 | 0 | 1 | 0.17 | 0.18 |

**Source:** Based on data from the Multiple Indicator Cluster Survey (MICS5) for the Dominican Republic (10).

To understand the extent to which variations of socioeconomic position co-occur with variations of other intervening factors in childhood development, we conducted a Pearson’s correlation analysis to explore: 1) the relationship between socioeconomic position and early childhood stimulation; and 2) the correlation between socioeconomic position and child discipline. We found significant positive correlations between socioeconomic position and early childhood stimulation: the availability of stimulating activities at home, with a small proportion of shared variance (*r*(7354) = .30, *p* < .001, *r*^*2*^ = .09); numbers of books at home, with a medium proportion of shared variance (*r*(7354) = .42, *p* < .0001, *r*^*2*^ = .18); and attendance to an early childhood education center, with a medium proportion of shared variance (*r*(7354) = .4, *p* <.001, *r*^*2*^ = .16). That is, children from more affluent homes have access to more stimulating resources and activities than children from less affluent homes. We found a significant negative correlation between socioeconomic position and the availability of stimulating objects at home, although the magnitude of the correlation was negligible (*r*(7354) = -.03, *p* = .017, *r*^*2*^ = .0008). Descriptive statistics are displayed in **Table 2**.

We found a positive significant correlation between socioeconomic position and the use of positive discipline by parents and caregivers, with a small proportion of shared variance (*r*(7354) = .12, *p* < .001, *r*^*2*^ = .02). This means that the higher the socioeconomic position, the higher the practice of positive child discipline, although we found no significant correlation between socioeconomic position and the use of negative discipline by parents and caregivers (*r*(7354) = -.02, p = .12). Descriptive statistics are displayed in **Table 2**.

We conducted a two-step hierarchical multiple regression analysis with ECD as the dependent variable. Step one corresponded to a sociodemographic model. We entered the independent variables: child’s age in months (SD1), child’s sex at birth (SD2), family socioeconomic position (SD3), and mother’s level of education (SD4). Step two corresponded to a sociodemographic + psychosocial model to which we added the psychosocial variables: stimulating activities at home (PS1), stimulating objects at home (PS2), the number of books at home (PS3), attendance to an early childhood education program (PS4), positive discipline at home (PS5), and negative discipline at home (PS6). Regression statistics are reported in **Table 3**.

**Table 3:**
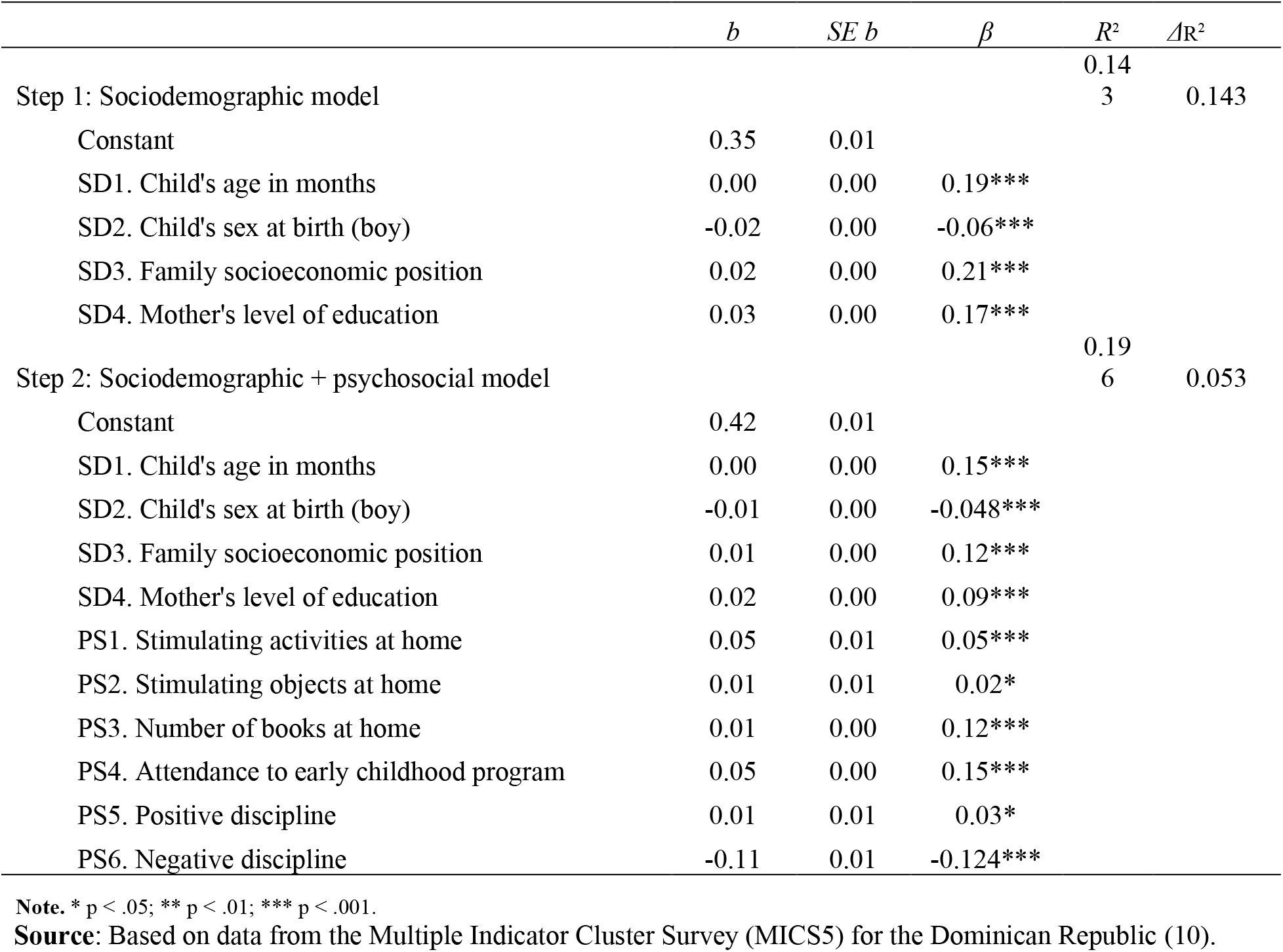
Summary of hierarchical regression analysis for variables predicting early childhood development.

Before conducting the hierarchical multiple regression analysis, we checked the linearity, normality, homoscedasticity, and multicollinearity assumptions for this particular test. The assumptions of linearity, normality, and homoscedasticity were met, as evidenced by the residual and scatter plots. Variance inflation factor statistics scored below 10 for all the variables, confirming that there was no multicollinearity.

The hierarchical multiple regression analysis showed that, at step one, the sociodemographic model contributed significantly to the regression model (*F*(4,3751) = 306.44, *p* < .001) and accounted for 14.3% of the variance in ECD. When introducing the psychosocial variables, the new model explained an additional 5.3% of the variance. This change in *R*^*2*^ was significant (*F*(6,7345) = 80.62, *p* < .001). Together, the sociodemographic and psychosocial variables accounted for 19.6% of the variance in ECD.

All regression coefficients were significant in both models. Most of the coefficients had positive valences, which means that they had positive effects on childhood development. However, two independent variables had negative regression coefficients: sex at birth and negative discipline. The results indicate a negative impact on ECD of being born male, which explained 6% and 4.8% of ECD variance in the sociodemographic and psychosocial models, respectively. Negative discipline negatively impacted child development, explaining 12.4% of the variance of ECD in the psychosocial model.

The most relevant sociodemographic predictors of ECD were socioeconomic position and mother’s education, which uniquely explained 21.0% and 17.0% of the ECD variance in the sociodemographic model, respectively. The most important psychosocial predictors of early childhood development were: 1) attendance to an early childhood education program, which uniquely explained 15.0% of the variance in ECD; 2) negative discipline, which uniquely explained 12.4% of the variance; 3) the number of children’s books at home, which uniquely explained 12.0% of the variance; and 4) stimulating activities at home, which uniquely explained 5% of the variance. **Figure 1** shows a visual representation that compares the standardized coefficients from both models. The variance explained by the sociodemographic + psychosocial model includes the predictive power of the factors that already belonged to the sociodemographic model. This is more evident for three factors: child’s age in months, family socioeconomic position, and mother’s level of education.

**Figure 1:**
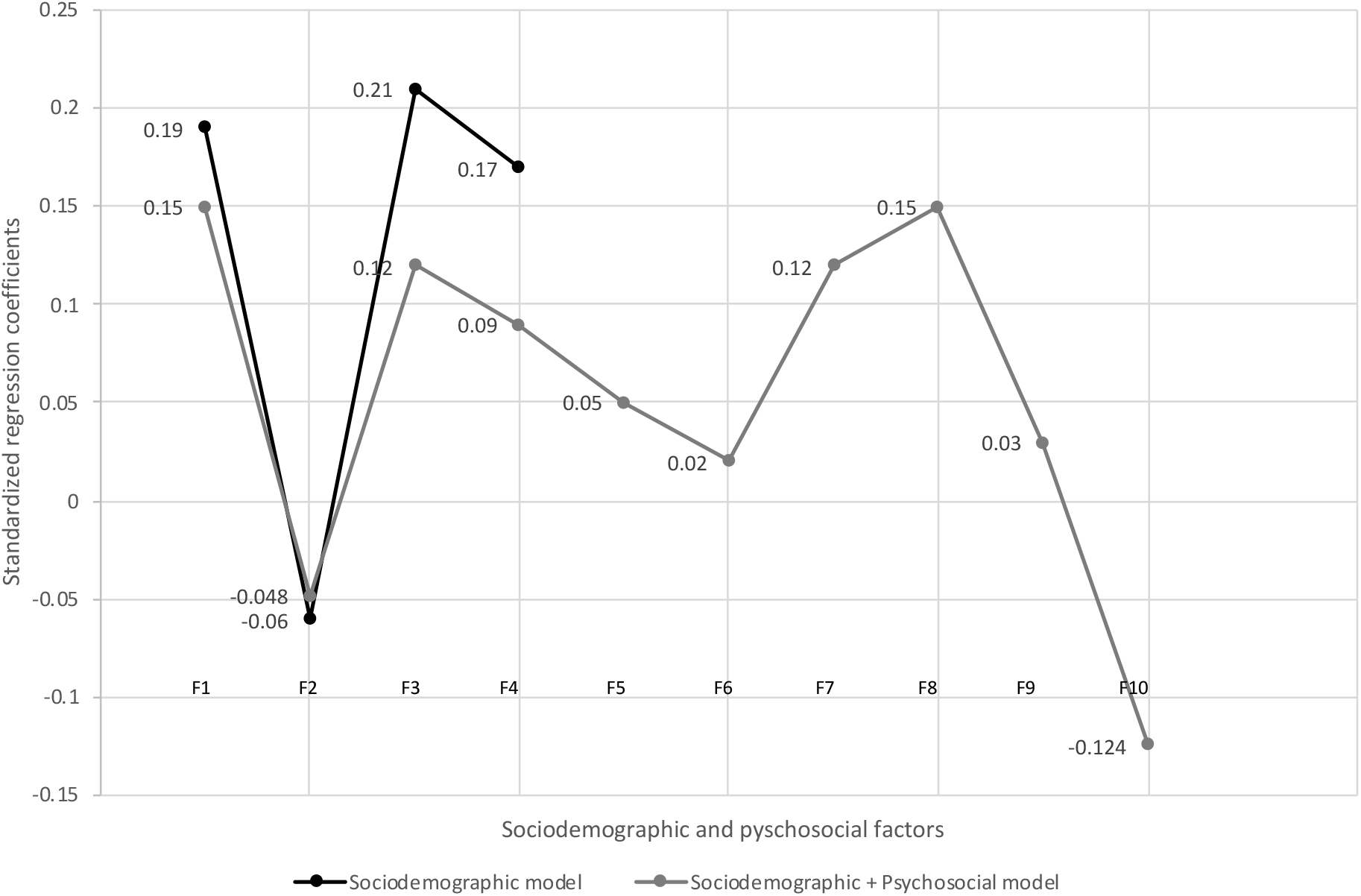
Regression coefficients for models that predict early childhood development. F1 = Child’s age in months; F2 = Child’s sex (boy); F3 = Family socioeconomic position; F4 = Mother’s level of education; F5 = Stimulating activities at home; F6 = Stimulating objects at home; F7 = Amount of books at home; F8 = Attendance to early childhood program; F9 = Positive discipline; F10 = Negative discipline.

Additionally, we explored the implications of the survey question that asked mothers or primary caregivers to indicate whether they identified with the following statement: “Children must be physically punished.” We divided our sample into two groups: caregivers who believed that physical punishment is necessary and those who believed that physical punishment is not necessary. We conducted a series of analyses of covariance (ANCOVA) to test differences between the groups on ECD, early childhood stimulation, and child discipline, controlling for socioeconomic position. We included socioeconomic position as a covariate given its correlation with most tested variables and to prevent confounding results. **Table 4** shows the descriptive statistics by group.

**Table 4:**
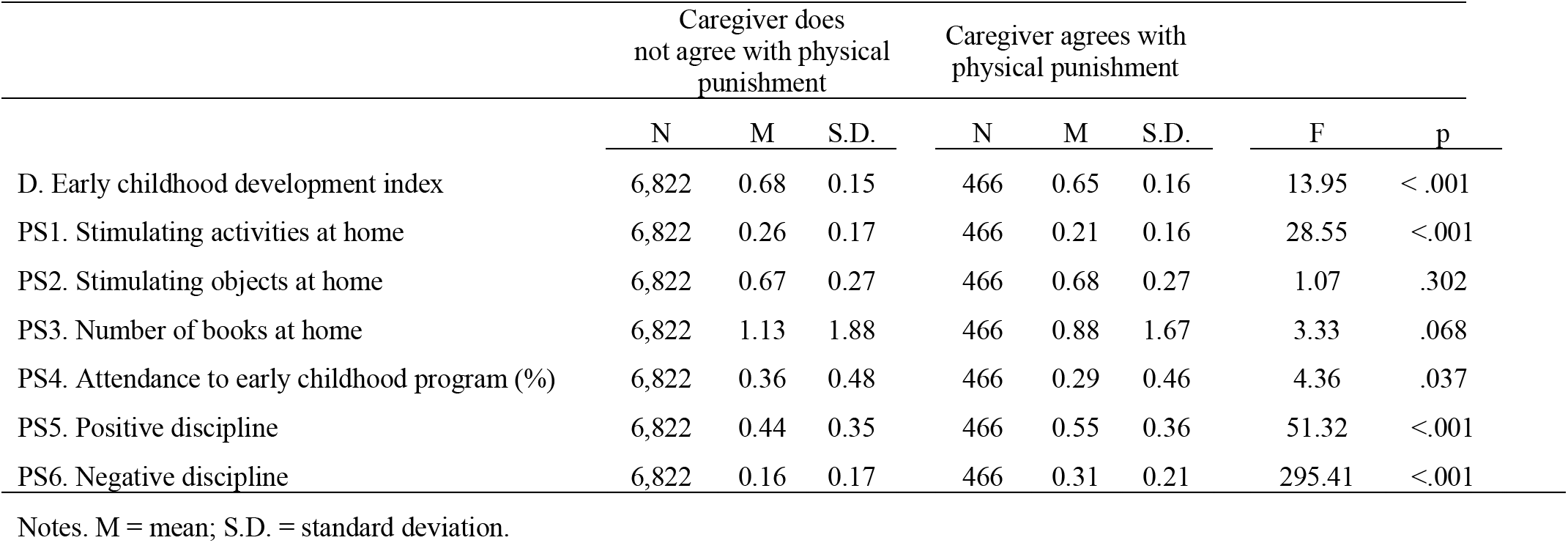
Descriptive statistics by caregivers who agree and who do not agree that children must be physically punished.

|  | Caregiver does not agree with physical punishment |  |  | Caregiver agrees with physical punishment |  |  | F | p |
| --- | --- | --- | --- | --- | --- | --- | --- | --- |
|  | N | M | S.D. | N | M | S.D. |  |  |
| D. Early childhood development index | 6,822 | 0.68 | 0.15 | 466 | 0.65 | 0.16 | 13.95 | < .001 |
| PS1. Stimulating activities at home | 6,822 | 0.26 | 0.17 | 466 | 0.21 | 0.16 | 28.55 | <.001 |
| PS2. Stimulating objects at home | 6,822 | 0.67 | 0.27 | 466 | 0.68 | 0.27 | 1.07 | .302 |
| PS3. Number of books at home | 6,822 | 1.13 | 1.88 | 466 | 0.88 | 1.67 | 3.33 | .068 |
| PS4. Attendance to early childhood program (%) | 6,822 | 0.36 | 0.48 | 466 | 0.29 | 0.46 | 4.36 | .037 |
| PS5. Positive discipline | 6,822 | 0.44 | 0.35 | 466 | 0.55 | 0.36 | 51.32 | <.001 |
| PS6. Negative discipline | 6,822 | 0.16 | 0.17 | 466 | 0.31 | 0.21 | 295.41 | <.001 |

Notes. M = mean; S.D. = standard deviation.

The results indicate that children whose caregivers do not agree with physical punishment compared with children whose caregivers agree with physical punishment, controlling for socioeconomic position, had: better development scores (*F*(1,7285) = 13.95, *p* < .001); higher attendance at a childhood education center (*F*(1,7285) = 4.36, *p* < .001); and had more availability of stimulating activities at home (*F*(1,7285) = 28.55, *p* < .001). There were no significant differences between the groups on the availability of stimulating objects and number of books at home, when controlling for socioeconomic position. Finally, caregivers who agree with physical punishment engage more both in positive discipline (*F*(1, 7285) = 51.32, p < .001) and negative discipline (*F*(1,7285) = 295.41, *p* < .001) compared with caregivers who do not agree with physical punishment, when controlling for socioeconomic position.

## Discussion

This study confirmed that the well-established notion that wealth and inequality impact childhood development is valid in the Dominican Republic. More specifically, we found that Dominican children from lower socioeconomic positions had lower childhood development scores than children from more affluent homes. We found that a predictive model that includes both sociodemographic (age, sex at birth, socioeconomic position, mother’s level of education) and psychosocial factors (childhood stimulation and child discipline) predict early childhood development above and beyond the sociodemographic factors alone. Psychosocial factors such as attendance to childhood education programs, availability of children’s books at home, and availability of stimulating activities at home were the strongest positive predictors of ECD. On the other hand, the use of negative discipline was the strongest negative predictor of childhood development. Therefore, psychosocial factors moderate the relationship between sociodemographic factors and childhood development. In other words, engaging in childhood stimulation—especially the attendance at childhood education programs, availability of books at home, and activities—and not using negative discipline buffer the effects of socioeconomic position and maternal education on childhood development.

Similar results have been found elsewhere. In a study with data from 26 low- and middle-income countries, literacy and numeracy were associated with attendance at an early childhood program, home literacy, and stimulating activities at home, and the effect of socioeconomic position on development was moderated by family care behavior (38). A study conducted by Tran et al. with data from 35 countries found that parental engagement in learning activities at home, hard punishment, and attendance to early childhood education moderate the effects of family poverty on development, but only in countries of low and medium human development index, and not in countries of high HDI (48). The Dominican Republic is categorized as of high HDI since 2010, four years before the MICS survey, but of medium HDI when adjusted for inequality (49). We hypothesize that this discrepancy could be due to the large role that inequality plays in preventing the poorest children from thriving in countries of high HDI but with a large share of the income concentrated within the wealthiest 10% of the population—35.4% in the Dominican Republic in 2010-17 (49).

This study found that positive discipline was more likely to be used by parents and caregivers from higher socioeconomic positions; however, negative discipline—one of the strongest factors that impact childhood development—was used regardless of socioeconomic position. Other studies have shown that negative discipline—specifically physical punishment—predicts young children’s aggressive behavior (41), which could then create a vicious cycle of aggressive childhood behavior that triggers aggressive discipline methods in adulthood. Lansford and Deater-Deckard suggested that there are country-specific factors associated with the use of violence to discipline children, evidenced by a large variability of the reported use of negative discipline in a study of 24 low- and middle-income countries in MICS household surveys (50). These sociocultural aspects should be studied in depth in the Dominican Republic, as 62.9% of children from 1 to 14 years of age had experienced negative violence during the month prior to the survey (10).

Our study also found that children whose caregivers believe in physical punishment had lower development scores. However, a few details should be pointed out: more children from caregivers who do not believe in physical punishment attend early childhood education programs compared to children from caregivers who believe in physical punishment (36% vs. 29%; p=0.037). There is some evidence that attendance to early childhood programs has a positive effect on children’s social competence, including behavior (51). This might make parenting easier and, therefore, more positive. Further studies should address children’s behavior at home and its relationship with attendance to early childhood programs to better understand this result. Also, early childhood programs usually offer interventions that target parenting skills (43), and this could explain part of the reasons why caregivers whose children attend early childhood programs do not agree with physical punishment as a discipline method.

Another interesting result was that caregivers who do not believe in physical punishment provide more stimulating activities at home than caregivers who believe in physical punishment (26% vs. 21%; p<.001), but not necessarily more books, toys, or stimulating objects. The actual social interaction within stimulating activities might improve children’s behavior, and not so much the available props with which to play. This finding supports Worku and colleagues’ work that reported moderate to large gains in social-emotional and language development from a home visit intervention that targeted mother-child play interaction (46). Finally, we found that caregivers who believed in physical punishment used more positive and also more negative discipline methods. Therefore, positive and negative discipline methods are not mutually exclusive, as has been shown earlier in Caribbean countries (52), and a caregiver of a child with challenging behaviors might use multiple discipline strategies.

This study has some limitations. One of them is a threat to internal validity due to instrumentation. First, the early childhood development instrument has poor internal consistency. This could be the result of ECD being a multidimensional construct. In fact, the instrument classifies early childhood development into four categories: literacy and numeracy development, physical development, social and emotional development, and approaches to learning. Some of these categories are measured with only two items, making it unsuitable for internal consistency testing. For example, the physical development subtest is measured by asking caregivers: a) if the child is too sick to play, and b) if the child can pick up a small object with two fingers. These two items are unrelated, and fewer items on a scale usually give lower scores. Notwithstanding the internal consistency limitation, this study’s tested models were strong enough to yield significant results, which means that the effects are seen even with a weak instrument.

Second, the instrument uses the same items to assess children ranging from 36 to 59 months of age. Childhood development is a continuum that changes quickly during the first month of life, and, therefore, a 59-month-old child will outperform a 36-month-old child on any development test. We overcame this limitation by adding the independent variable age in months to the sociodemographic model, to account for differences across age groups. Third, there are country-specific characteristics with which this instrument might interfere. For example, the instrument item “can read simple common words” is part of the developmental assessment. Still, in the Dominican Republic, the official literacy training occurs in first grade, when children are six years old, which is older than the MICS5 age group. This issue, reported elsewhere (53), places most Dominican children at risk of underscoring when the reality is that they have not been trained to respond to such stimuli just yet. However, it is important to mention that this instrument belongs to a household survey, whose intention is not to provide a comprehensive assessment of ECD, but as a monitoring tool to evidence broad progress and allow cross-country comparisons.

Third, the Dominican Republic does not have a standardized instrument to screen for ECD. Anecdotally, many institutions report to have used the Denver Developmental Screening Test II (54) and other self-made assessments, but systematic data collection has yet to occur. An instrument under consideration to use at a national scale for this purpose is the Dominican Screener for Childhood Development (55), an adaptation of the Malawi Developmental Assessment Tool (56).

Fourth, evidence from various settings has demonstrated that malnutrition, including micronutrient deficiencies, may prevent optimal development among children and adolescents (57-60). However, we were unable to add nutrition to the models because MICS5 only reports nutrition variables for children ages 0 to 23 months old. To our knowledge, no research has specifically examined this phenomenon in the Dominican Republic, where stunting has decreased among children under 5 years from 19.4% in 1991 to 6.9% in 2013 but is distributed unequally between children from the poorest wealth quintile (11.3%) and those from the richest (3.9%) (61, 62). Additionally, household survey data from 2014 show that more than half of children aged 6 to 23 months did not have a minimum acceptable diet (10)—in terms of nutritional diversity or frequency of meals as defined by UNICEF (63).

Notwithstanding these limitations, this study is relevant because it provides a better understanding of some specific characteristics of how childrearing in the Dominican Republic is associated with ECD and what types of activities should be prioritized to prevent childhood developmental delay. In recent years, the Dominican Republic has made enormous efforts to promote ECD, such as the creation in 2015 of the National Plan for Early Childhood Comprehensive Protection and Care and the National Institute for Early Childhood Comprehensive Care (INAIPI, for its acronym in Spanish) (64), which offers home visit programs and early childhood education centers in the areas of highest social vulnerability at no cost. Although the coverage does not yet reach the entire target population, it is an indication that the country is making progress. For multisector approaches to be effective, they need to consider and integrate the social, economic, political, and cultural contexts, the environment for caregivers, and the types of nurturing care (health, nutrition, security and safety, responsive caregiving, and early learning) (65)—in such ways that they promote interventions that mitigate the multiple risk factors for developmental delays and that promote nurturing caregiving, including socio-emotional and cognitive stimulation (66). Although the results of this study show a protective effect of psychosocial factors, sustainable and large-scale intervention should not be limited to just buffering, but to actually solve the underlying problem which is that poverty prevents children from reaching their developmental potential.

## Data Availability

This study used de-identified secondary data that are publicly available.

http://mics.unicef.org

## Acknowledgements

We thank Carlos Ruiz-Matuk for his support during data analysis and Virginia Savage and Liz Tracy for assisting with literature review.

## Sources of support

The study was funded by the 2018 Carol Lavin Bernick Faculty Grant Program at Tulane University made to Arachu Castro, who was also funded through gifts from the Zemurray Foundation for her position as the Samuel Z. Stone Chair of Public Health in Latin America at the Tulane School of Public Health and Tropical Medicine (New Orleans, Louisiana, United States).

## Notes

### Competing Interest Statement

The authors have declared no competing interest.

### Author Declarations

After registration at http://mics.unicef.org/ by entering a username and password and requesting access to conduct secondary data analysis of childhood development indicators, the UNICEF MICS team approved access to the MICS datasets on February 19, 2018. The MICS survey responds to local regulations and protocols regarding data collection and fieldwork (CITE). The Dominican Republic does not demand to obtain approval from an Ethical review board. However, the MICS protocol requires all data to be kept strictly confidential, including secure storage of records and databases with no identifiers. In addition, the protocol requires the interviewer to obtain appropriate informed consent from survey respondents. More information available in the MICS Manual, pages 25-26, at: https://mics.unicef.org/files?job=W1siZiIsIjIwMTUvMDMvMzEvMDgvNTUvMjYvOTg3L01JQ1M0X01hbnVhbF9QcmVwYXJpbmdfZm9yX0RhdGFfQ29sbGVjdGlvbl9hbmRfQ29uZHVjdGluZ19GaWVsZHdvcmsuZG9jeCJdXQ&;sha=8d74d95f24a423eb.

